# The Third Dimension of Pharmacokinetic–Pharmacodynamic Theory: Adaptive Rate Capacity as a Conserved Constraint on Biological Tolerability

**DOI:** 10.64898/2026.06.02.26354717

**Authors:** Cornelis H Kleinbloesem, Cornelis L Braal

## Abstract

Pharmacokinetic–pharmacodynamic theory describes drug action in two dimensions, magnitude and time, yet rate-dependent toxicity recurs across unrelated therapies without a unifying principle. We propose the Human Adaptive Rate Limit (HARL): every physiological system tolerates a maximum velocity of state change, |dS/dt|max = σmax/τ, set by its reserve σ and adaptation time τ. We show that nifedipine, erythropoiesis-stimulating agents, radiation, vancomycin and CAR T-cell therapy obey one velocity boundary spanning minutes to weeks. Reanalysing the 202-patient CAR-T cohort of Wei et al., a dual-velocity classifier (ferritin and D-dimer rate of rise) predicted severe cytokine release syndrome with 100% sensitivity and a median 4-day lead time before clinical onset. Velocity is a third, governable dimension of tolerability; a dynamic maximum tolerated rate should complement the maximum tolerated dose in drug development.

---

Pharmacokinetic–pharmacodynamic modelling has achieved remarkable sophistication over four decades.^1–3^ Population approaches, mechanism-based models, and physiologically-based frameworks routinely guide dose selection. Yet classical PK/PD operates within a fundamentally two-dimensional parameter space: how much drug is present and how the system responds over time. What this framework lacks is formal representation of the rate at which biological systems can adapt to pharmacologically-driven state changes.

This absence matters because, for drugs engaging adaptive systems—vascular remodelling, erythropoietic expansion, immunological checkpoints—trajectory determines tolerability as much as destination. Two patients reaching identical pharmacodynamic states can experience profoundly different outcomes depending on whether adaptive machinery has kept pace. Dose and exposure are scalars; harm in adaptive systems is frequently a derivative: uncontrolled velocity of state change.^4,5^

The need for formalisation has become urgent. Therapeutic modalities increasingly deploy rapid-acting biological forcing functions—bispecific engagers, CAR-T cells, mRNA therapeutics, gene editing—that drive state changes faster than classical small molecules.^6–9^ Step-up dosing, mandated hospitalisation, and infusion-rate restrictions represent empirical recognition of velocity governance, yet no quantitative framework connects these domain-specific practices.

Here we formalise the Human Adaptive Rate Limit (HARL) and demonstrate its operation across five therapeutic domains. Foundational evidence from a prospective crossover study that causally isolated rate of rise as a tolerability variable^10^ predicted a population-level mortality signal subsequently confirmed.^11,12^ We show that the same constraint explains rate-dependent toxicity in haematology, radiation, infusion pharmacology, and immunotherapy, and that velocity-based surveillance predicts severe cytokine release syndrome with clinically actionable lead time.

## The State-Adaptation Mismatch framework

Let S(t) denote the pharmacodynamically-driven system state and A(t) the adaptive trajectory the organism executes without injury. The stress gap is σ(t) = |S(t) − A(t)|. Adaptation operates on characteristic timescales (τ) set by biology: weeks for vascular remodelling (τ ≈ 14–28 days), days for immunoregulatory mechanisms (τ ≈ 2–7 days), hours for DNA repair (τ ≈ 0.5–12 h), minutes for histamine degranulation buffering (τ ≈ 2–10 min). Adaptation follows first-order kinetics:

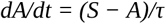

Toxicity emerges when σ > σ^max^. Under constant forcing velocity v = dS/dt, the steady-state gap is σ^ss^ = v.τ. Requiring σ^ss^ ≤ σ^max^ yields the velocity limit:

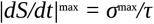

This is the HARL—a universal velocity boundary (figure 1). Systems with long τ tolerate only slow changes; those with short τ accommodate faster perturbations but remain bounded. For non-constant forcing, the general convolution solution confirms the bound for arbitrary waveforms (web appendix pp 1– 2). The clinical consequence: dosing protocols should specify both target state and allowable rate of approach—Maximum Tolerated Dynamics (MTDyn).

**Figure 1:**
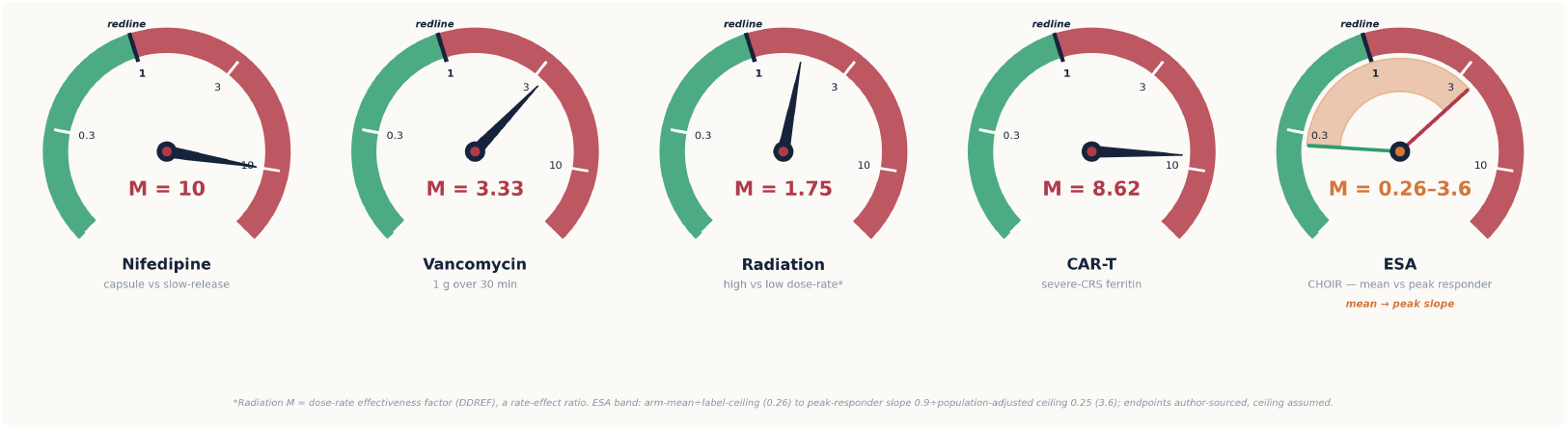
The velocity speedometer. Each domain’s measured rate of state change is read against its own adaptive ceiling σ/τ, giving the dimensionless mismatch M (redline at M = 1). Four domains breach—nifedipine ≈10, CAR-T 8.6, vancomycin 3.3, radiation 1.8 (dose-rate effectiveness factor)—while ESA spans the redline: safe on the trial arm-mean rate (M = 0.26) but breaching on the peak-responder slope (M = 3.6). Velocity must be read as a peak, not an average.

### Foundational evidence: nifedipine (1984–1996)

The empirical foundation comes from a prospective crossover study designed to isolate pharmacokinetic input rate as a causal variable.^10^ Eight healthy volunteers received nifedipine via computer-controlled infusion pump under two protocols matched for steady-state concentration (C ^ss^ ≈ 35 ng/mL): a gradual 4-h ramp versus rapid attainment within 30 min followed by deliberate acceleration at steady state. By driving subjects to identical concentrations via different trajectories, the design isolates dC/dt as the sole experimental variable.

Rapid approach segments produced sustained reflex tachycardia (ΔHR +15–25 bpm) absent in slow-rise regimens, despite equivalent blood pressure reduction.^10^ The baroreflex accommodation timescale (τ ≈ minutes) was outrun by the rapid input, producing autonomic decompensation at ‘acceptable’ plasma levels. Critically, the formulation-dependent divergence in heart rate, blood pressure, and plasma noradrenaline was first reported in 1984^14^—before any regulatory reaction—with the full pharmacokinetic– pharmacodynamic relationship characterised in the same period^13^. The 1987 controlled-infusion study isolated rate of rise as the causal variable; the dose-related population mortality signal and the consequent clinical advisories followed only in 1995.

This finding generated a testable prediction: formulations permitting rapid systemic input should produce excess cardiovascular harm at population scale. Furberg et al (1995) confirmed this in ∼8350 patients with coronary heart disease, reporting dose-related excess mortality with short-acting nifedipine.^11^ The signal concentrated in patients with impaired coronary reserve—reduced σ^max^ for haemodynamic perturbation. Grossman et al (1996) concluded in JAMA that sublingual nifedipine for hypertensive urgencies posed unacceptable risk, catalysing a de facto clinical moratorium.^12^ This arc—mechanistic isolation → later population signal → policy change—shows the framework can anticipate harm rather than merely rationalise it post hoc (figure 2).

**Figure 2:**
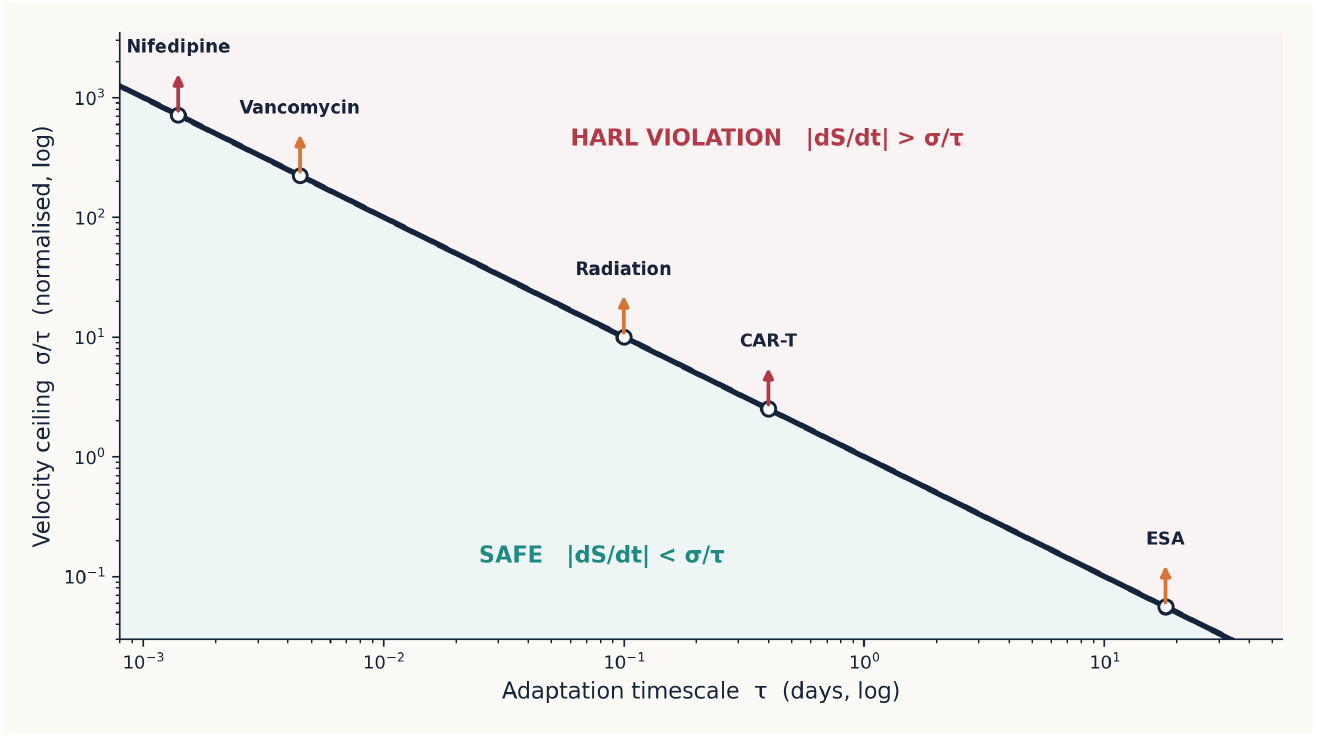
The velocity envelope. The maximum tolerable velocity |dS/dt|max = σ/τ falls as a system’s adaptation time τ lengthens (σ normalised to 1). Each domain is placed by its τ; arrows mark that the measured velocity exceeds its own ceiling. Axes are velocity and its limit only—no cost—so the boundary is comparable across domains spanning minutes (vancomycin, nifedipine) to weeks (ESA).

### Convergent evidence across therapeutic domains

If HARL reflects a general biological principle, it should manifest wherever pharmacological forcing engages adaptive systems with finite rate capacity. We present four additional domains with distinct mechanisms but the same velocity boundary.

#### Erythropoiesis-stimulating agents

The CHOIR trial randomly assigned 1432 patients with chronic kidney disease (eGFR 15–50 mL/min) to aggressive (target Hb 13.5 g/dL) versus conservative (11.3 g/dL) correction with epoetin alfa. ^15^ The aggressive arm produced significantly more cardiovascular events: hazard ratio (HR) 1.34 (95% CI 1.03– 1.74; p=0.03), 125 events (17.5%) versus 97 (13.5%), number needed to harm 25. The DSMB terminated the trial early.

Three trials corroborated this. CREATE showed trends toward harm with full normalisation and higher dialysis initiation.^16^ TREAT demonstrated twofold stroke risk with darbepoetin (HR 1.92) in diabetic CKD.^17^ The Normal Hematocrit Trial was terminated for excess mortality targeting haematocrit 42%.^18^ Interpreting CHOIR through HARL requires care. At the trial arm-mean rate of rise the velocity ceiling was respected (M≈0.26), so the excess events are substantially a level effect rather than a velocity one. The rate signal emerges only in the rapid-responder subgroup, whose peak slope can exceed a population-adjusted ceiling (M up to ∼3.6); secondary analyses are consistent with worse outcomes in faster responders, and ESA dose increments were independently associated with events. ESA therefore sits at the boundary—the negative-control domain in which the velocity axis is engaged only under aggressive titration, and in which mean-based rates can average the breach away.^19^

In SAM terms, rapid Hb correction in uraemic vessels with impaired compliance outpaces vascular adaptation (τ ≈ days to weeks). Viscosity rises before plasma volume adjusts; shear stress increases before endothelial remodelling; prothrombotic risk develops before haemostatic rebalancing. Current FDA prescribing embeds velocity governance: target Hb 10–11 g/dL, rate ≤1 g/dL per 2 weeks—de facto MTDyn (figure 3).^20^

**Figure 3:**
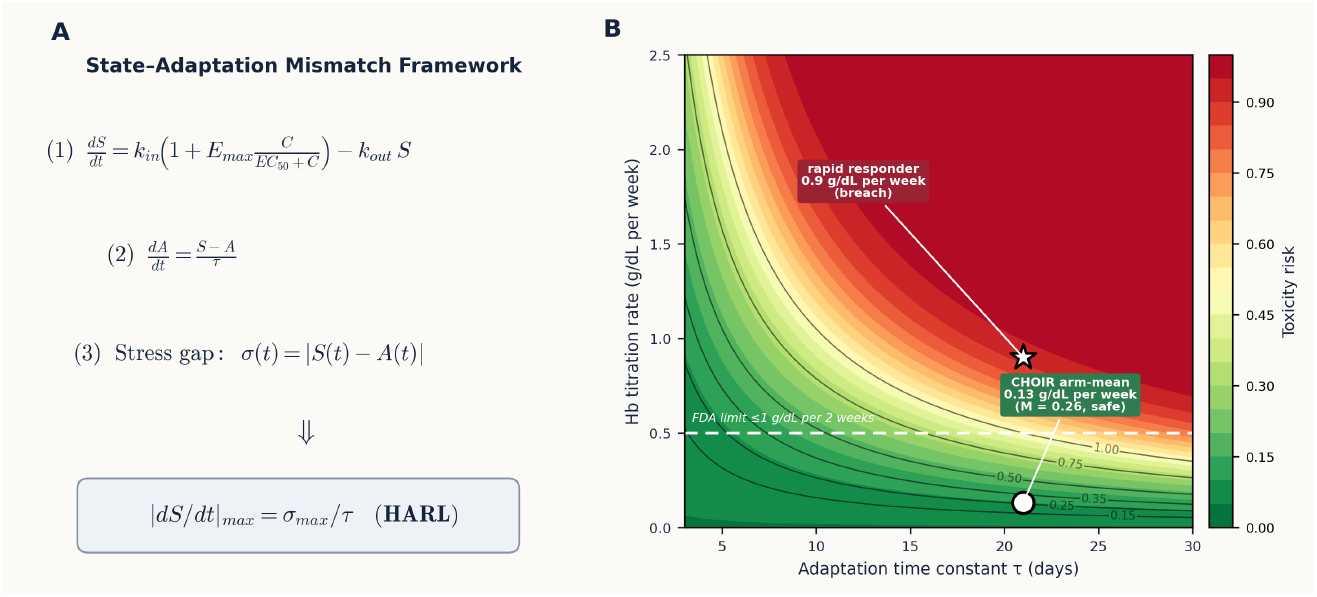
The SAM framework and the ESA negative control. (A) State-Adaptation Mismatch framework: equations for forcing S(t), adaptive response A(t), and stress gap σ(t); the HARL boundary |dS/dt|max = σmax/τ follows directly. (B) Toxicity-risk surface for ESA therapy as a function of adaptation time constant τ and haemoglobin titration rate. The CHOIR arm-mean rate (0.13 g/dL per week, circle) sits in the low-risk green zone below the FDA rate limit (M = 0.26); only the rapid-responder subgroup (0.9 g/dL per week, star) crosses into the high-risk zone. ESA is therefore the negative-control domain: the mean rate respects the ceiling, and the velocity signal appears only under aggressive titration.

#### Radiation biology

Radiation provides the most quantitatively precise demonstration of dose-rate dependence. Acute whole-body delivery of 10 Gy is uniformly lethal (LD^50/60^ ≈ 4–5 Gy; 10 Gy produces inevitable gastrointestinal syndrome within 2–3 weeks).^21^ The identical 10 Gy fractionated over weeks (2 Gy/fraction, 5 fractions/week) is routine curative treatment. The dose-rate effectiveness factor (DDREF 1.5–2.0, ICRP/BEIR VII) quantifies this: identical doses are 50–100% more damaging at high delivery rates.^22,23^

The mechanism is DNA repair kinetics. At >1 Gy/min, double-strand breaks accumulate faster than base excision repair, nucleotide excision repair, and homologous recombination can restore integrity. At <0.1 Gy/h, repair operates between hits (τ^repair^ ≈ 0.5–4 h). The five Rs of radiobiology—repair, redistribution, reoxygenation, repopulation, radiosensitivity—are time-dependent processes that HARL formalises under a single velocity constraint.^21^

#### Infusion pharmacology

Red Man syndrome—non-IgE histamine release with flushing, erythema, pruritus, and occasional cardiovascular collapse—demonstrates that infusion rate, independent of total dose, determines acute tolerability.^24,25^ Identical vancomycin doses cause reactions when infused rapidly but are tolerated over ≥60 min. Consensus guidelines specify velocity constraints: ≤10 mg/min; extend to 90–120 min above 1500 mg.^24^ The mast cell degranulation buffering timescale (τ ≈ 2–10 min) sets the HARL corridor. Vancomycin prescribing already implements MTDyn without invoking the theoretical framework.

### Immunotherapy: velocity-governed cytokine release syndrome

The preceding domains demonstrate HARL on timescales from minutes to weeks. CRS following CAR-T or bispecific antibody therapy engages the immune system on intermediate timescales (hours to days), providing the most detailed empirical validation through longitudinal biomarker data.^26–28^

CRS severity does not correlate simply with peak cytokine concentrations. The ASTCT consensus grades severity by downstream end-organ dysfunction—fever, hypotension, hypoxia, vasopressor requirements.^26^ By the time these thresholds are crossed, the inflammatory system has already transitioned into self-amplifying pathological escalation. This motivates a kinetic approach: can the *velocity* of inflammatory acceleration identify patients destined for severe CRS before magnitude criteria declare storm?

The mechanistic basis is a staged cascade (figure 4). Tumour engagement (hours 0–24) triggers macrophage activation with ferritin velocity as the first signal (days 1–3). Activated macrophages produce IL-6, IL-1β, TNF-α in a positive feedback loop (days 2–5). Sustained cytokines activate endothelial and coagulation cascades with D-dimer velocity as the downstream signal (days 3–7) before end-organ dysfunction (days 5– 10).^29–32^ Bispecific engagers (teclistamab, glofitamab, epcoritamab) now mandate step-up dosing with hospitalisation—explicit velocity governance.^33–35^

**Figure 4:**
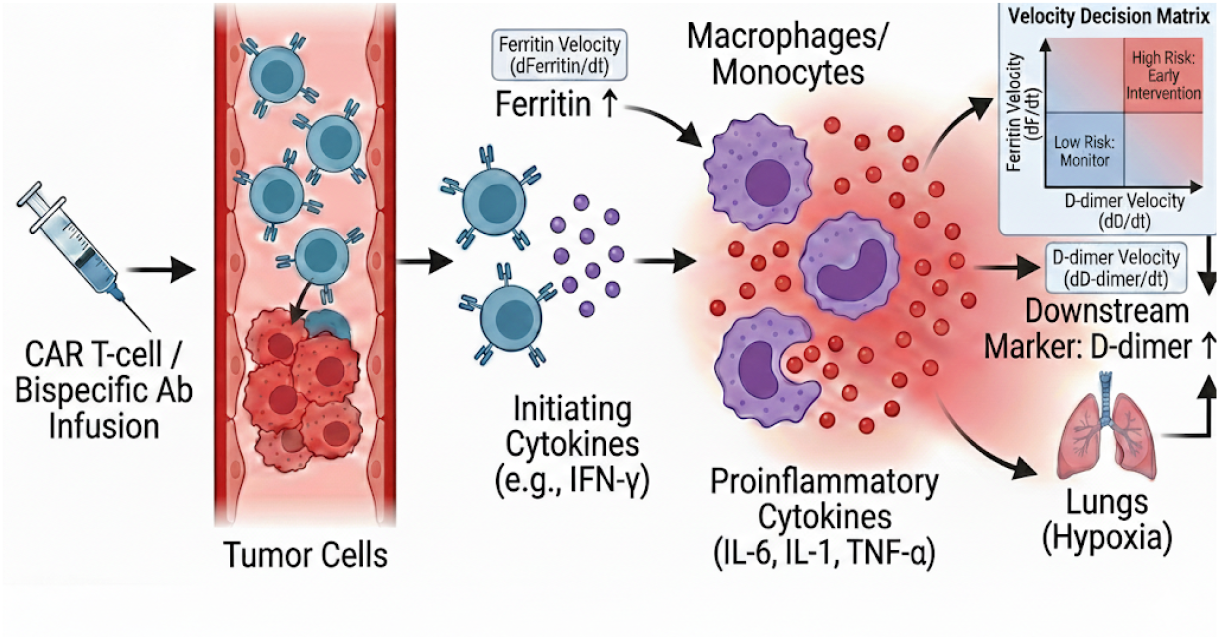
CRS inflammatory cascade. Progression from CAR-T infusion through tumour engagement, macrophage activation (ferritin velocity), cytokine amplification, endothelial activation (D-dimer velocity), to organ dysfunction. Velocity decision matrix (upper right) shows early kinetic risk stratification.

### Empirical validation: velocity-based CRS prediction (N=202)

We reanalysed the longitudinal biomarker data from the 202-patient CAR-T cohort of Wei et al (2023), substituting biomarker velocity for the peak-level variables used in the original report.^36,37^ CRS was graded per ASTCT consensus; 45 patients (22.3%) developed grade ≥3.^26^

#### Performance

Two operating modes were evaluated on velocity thresholds of ferritin >232 ng/mL per day and D-dimer >1.21 mg/L per day. The safety rule-out mode, tuned for 100% sensitivity (miss no severe CRS), achieved specificity ∼78%. The Youden-optimised balanced mode achieved sensitivity 91.1% (95% CI 79.4–97.2), specificity 93.6% (95% CI 88.9–96.9), and AUC 0.95 (95% CI 0.91–0.98). Median kinetic lead time from velocity threshold crossing to clinical grade ≥3 CRS was 4 days (range 3–7; figure 5), providing a rational window for pre-emptive tocilizumab (IL-6 receptor antagonist) or corticosteroid intervention (figure 6). ^37^

**Figure 5:**
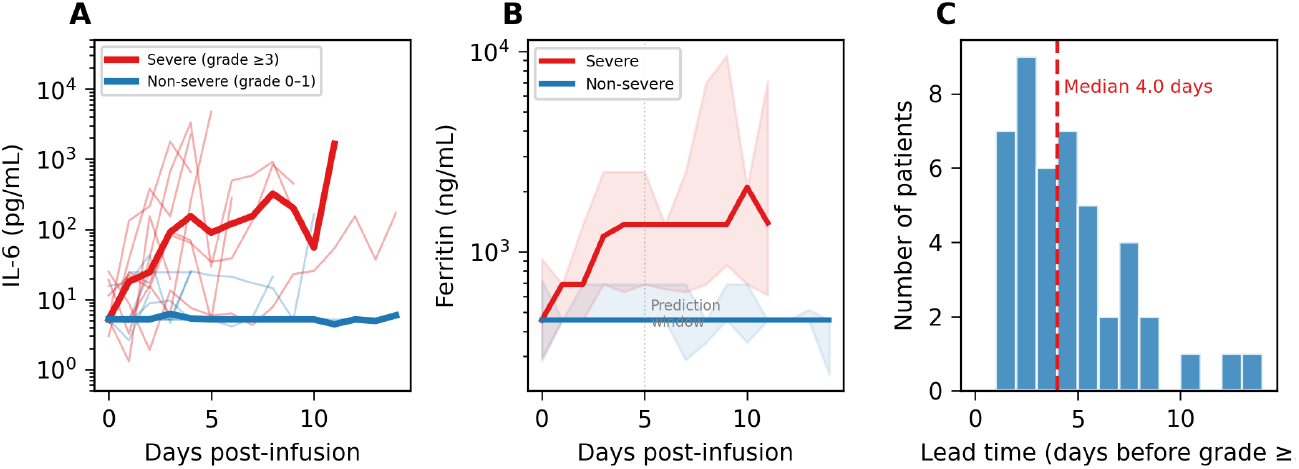
Real-data biomarker trajectories (N=202). (A) IL-6 trajectories (individual and medians) for severe (red) versus non-severe (blue) CRS; kinetic divergence by day 2–3. (B) Ferritin trajectories (medians, IQR). Dashed line: day 5 window. (C) Lead time distribution among severe cases.

**Figure 6:**
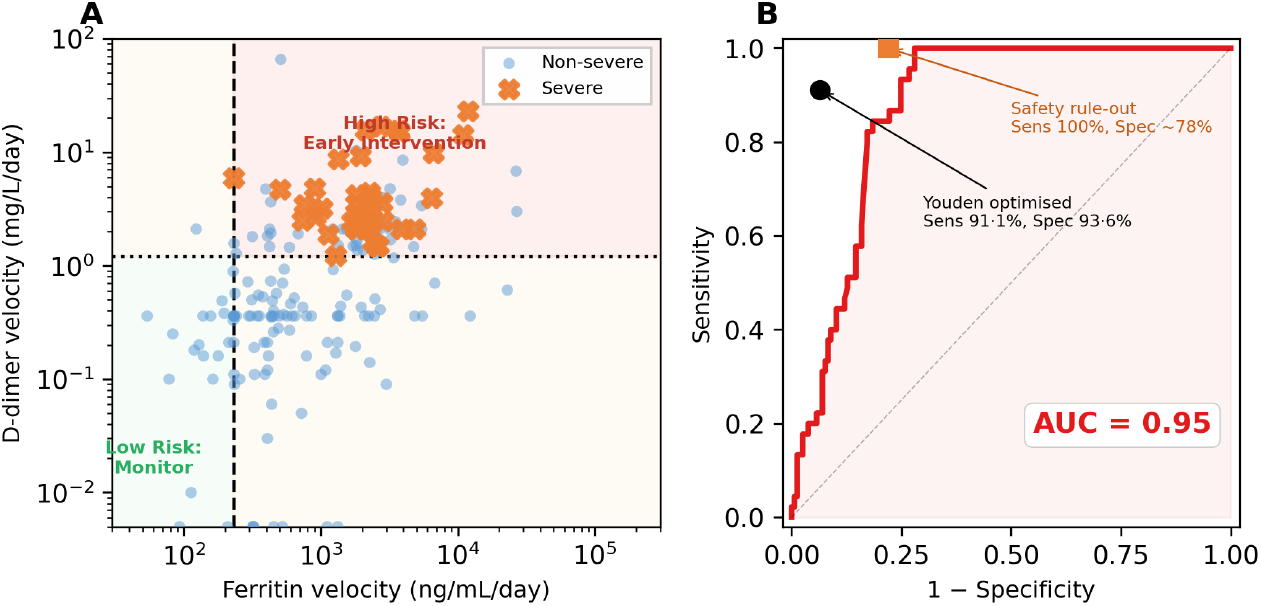
Velocity-based CRS prediction. Individual-patient kinetic decision matrix. Each point: one patient. Axes: maximum positive ferritin slope (x, ng/mL per day, log10) and maximum positive D-dimer slope (y, mg/L per day, log_10_) over days 0–5. Severe CRS: ASTCT grade ≥3. Blue circles: non-severe (n=157); orange crosses: severe (n=45). Vertical dashed: ferritin velocity threshold (232 ng/mL per day). Horizontal dotted: D-dimer velocity threshold (1.21 mg/L per day). Quadrant logic: patients are classified as high-risk when both ferritin AND D-dimer velocities exceed their respective thresholds (upper-right quadrant). All 45 severe patients occupy the upper-right quadrant. Sensitivity 100%, specificity ∼78%. (B) Receiver operating characteristic curve for the combined log-score classifier; AUC 0.95 (95% CI 0.91–0.98). Black circle: Youden-optimised operating point (sensitivity 91.1%, specificity 93.6%). Orange square: safety rule-out operating point (sensitivity 100%, specificity ∼78%).

### Cross-domain synthesis

Supplementary Table 1 summarises the empirically determined HARL parameters. For each domain, the state variable S (the pharmacodynamically-driven quantity whose velocity is rate-limited) is specified: heart rate for nifedipine, haemoglobin concentration for ESA, DNA damage burden for radiation, plasma histamine for vancomycin, and composite inflammatory activation (ferritin/IL-6/D-dimer) for immunotherapy. The inverse τ–velocity relationship across domains with fundamentally different state variables and molecular mechanisms supports HARL as a universal boundary condition.

## Discussion

This study establishes that biological tolerability is governed by three dimensions: exposure magnitude, time, and velocity of state change. A single equation—|dS/dt|^max^ = σ^max^/τ—unifies rate-dependent toxicity across five domains, three decades, and timescales from minutes to weeks. The nifedipine validation arc demonstrates predictive power: the 1987 study generated a falsifiable prediction confirmed in ∼8350 patients eight years later.^10–12^

The velocity-governed CRS prediction converts a reactive emergency into a prospectively monitorable process. The velocity classifier (sensitivity 100%, specificity ∼78%) and 3–7-day lead time provide a clinically meaningful intervention window for tocilizumab (IL-6 receptor antagonist), corticosteroids, and supportive care. This is proactive governance, not reactive rescue.^28,38,39^ The two-stage classifier mirrors the biological cascade: ferritin velocity as upstream macrophage signal, D-dimer velocity as downstream endothelial injury signal, enabling mechanistically interpretable risk stratification.

Prospective validation in an independent cohort is essential before clinical implementation. The safety rule-out specificity of ∼78% means that approximately one in five flagged patients will not develop severe CRS —an acceptable trade-off given the catastrophic consequence of missing decompensation and the low cost of heightened surveillance. The Youden-optimised mode (AUC 0.95) provides further risk stratification within the flagged population, enabling graded intervention intensity. Crucially, 100% sensitivity in this dataset is an empirical operating characteristic, not a guaranteed clinical property; external validation in independent cohorts with standardised sampling intervals is essential before clinical deployment.

HARL has implications for precision medicine beyond CRS. Adaptive capacity varies: elderly patients have reduced vascular compliance and slower receptor regulation (lower σ^max^, longer τ); cardiovascular disease impairs endothelial accommodation; high tumour burden amplifies immune forcing velocity. Personalising *tempo*—adjusting titration rates and fractionation to match individual adaptive reserves—extends precision medicine from dose individualisation to trajectory individualisation.^40,41^

For drug development, Phase I trials should establish MTDyn alongside MTD: dose-escalation with concurrent velocity-escalation arms. Formulation should be recognised as a safety mechanism that shapes the forcing function, not mere packaging. Regulatory agencies should require MTDyn characterisation for programmable therapeutics—CAR-T with safety switches, mRNA with tunable expression, fractionated gene editing—where velocity governance can be engineered at the design stage.^42–46^

### Panel 1: Emerging therapeutic modalities where HARL governance is critical

The HARL framework applies with particular urgency to next-generation therapeutic modalities that impose rapid, high-amplitude biological forcing functions. Unlike conventional small molecules—where absorption and distribution impose natural rate-limiting steps—engineered biological therapeutics can drive state changes at velocities that overwhelm adaptive systems within hours.

#### CAR-T and engineered cellular therapies

Current CAR-T products (tisagenlecleucel, axicabtagene ciloleucel, brexucabtagene, lisocabtagene, idecabtagene) drive exponential in-vivo expansion with peak forcing velocities determined by tumour burden, costimulatory domain (CD28 vs 4-1BB), and manufacturing variables. Next-generation constructs introduce additional rate-governing complexity: armoured CARs secreting cytokines (IL-12, IL-15, IL-18) add paracrine forcing on top of direct cytotoxicity; logic-gated CARs (synNotch, SUPRA-CAR) gate activation but may produce abrupt on/off switching rather than graded engagement; dual-targeting CARs (CD19/CD22, BCMA/CD38) multiply the engagement surface, amplifying the forcing function.

#### CAR-NK cells

Natural killer cells engineered with chimeric antigen receptors offer a different HARL profile: shorter in-vivo persistence (τ ≈ 7–14 days vs months for CAR-T), lower-amplitude but more rapid cytokine release (IL-15-armoured CAR-NK produce immediate IFN-γ and TNF-α forcing), and distinct innate immune activation pathways. The velocity constraint differs from CAR-T: peak forcing occurs earlier (hours 6–48 vs days 2–7) but at lower magnitude, meaning σ_max may be crossed transiently rather than sustained. Allogeneic off-the-shelf CAR-NK products (e.g. FT596, NKX101) add complement-mediated and host-vs-graft dynamics to the forcing function. HARL monitoring must capture these faster kinetics with higher-frequency biomarker sampling (≤6-h intervals).

#### CRISPR and in-vivo gene editing

Lipid nanoparticle-delivered CRISPR–Cas9 (e.g. NTLA-2001 for transthyretin amyloidosis) produces a fundamentally different forcing profile: a single bolus creates irreversible genomic change. The velocity of editing is determined by LNP biodistribution kinetics (τ_delivery ≈ hours), sgRNA–Cas9 complex formation (τ_editing ≈ minutes to hours), and cellular response to double-strand breaks (τ_repair ≈ hours to days). Off-target editing probability scales with Cas9 residence time, creating a dose-rate paradox: faster delivery increases on-target efficiency but also increases off-target risk per unit time. HARL governance requires titrating LNP dose and infusion rate to stay within the editing velocity corridor.

#### mRNA therapeutics

mRNA vaccines and therapeutics (mRNA-1273, BNT162b2, mRNA-based IL-12 intratumoral, personalised neoantigen vaccines) produce transient but intense protein expression. Translation velocity peaks 4–12 h post-injection with expression kinetics governed by LNP formulation, mRNA stability modifications (N1-methylpseudouridine, cap analogues), and codon optimisation. The innate immune response to mRNA itself—via TLR7/8, RIG-I, MDA5—adds a parallel forcing function independent of the encoded protein. Reactogenicity (fever, myalgia, fatigue) correlates with innate activation velocity rather than total antigen production. Self-amplifying RNA (saRNA) extends the expression window but may produce secondary velocity peaks as replicon-driven translation reignites immune activation days after initial dosing.

### Panel 2 (Web Appendix): Longevity gene editing—HARL under zero-tolerance constraints

Therapeutic gene editing for age-related disease and longevity applications represents the most stringent test case for HARL governance. Unlike oncology—where some toxicity is accepted against lethal disease—longevity interventions target otherwise healthy individuals with decades of remaining life expectancy. The acceptable σ_max approaches zero: any breach of the adaptive boundary produces net harm in a population with no disease-driven mortality to offset.

#### The irreversibility constraint

Gene editing is the only therapeutic modality where the pharmacodynamic effect is permanently encoded in the genome. A velocity violation in conventional pharmacology is self-correcting (drug clears, receptor recovers); in gene editing, any off-target modification persists through every subsequent cell division. The HARL framework must therefore account for cumulative velocity: total edits per cell per unit time across all targeted loci, including mosaicism patterns where some cells receive multiple edits while others receive none.

#### Candidate longevity targets and their HARL profiles

Telomerase activation (hTERT): Requires precise velocity—too slow produces insufficient telomere extension; too fast risks oncogenic transformation. The τ for telomere-mediated senescence reversal is weeks to months; the σ_max for neoplastic transformation is unknown but catastrophic. Klotho upregulation: Soluble α-Klotho modulates FGF23 signalling, Wnt pathways, and oxidative stress. Rapid systemic Klotho elevation could produce hypophosphataemia (by FGF23 suppression), disrupted calcium homeostasis, and paradoxical Wnt-driven proliferation in susceptible tissues. Follistatin / myostatin blockade: Rapid muscle hypertrophy via myostatin inhibition has produced cardiac hypertrophy in animal models when forcing velocity exceeds the cardiac remodelling timescale (τ_cardiac ≈ weeks). PCSK9 knockdown: VERVE-101 (base editing of PCSK9) aims for permanent LDL reduction. The velocity concern is hepatic: rapid, high-efficiency editing produces acute hepatocyte stress from simultaneous DSB repair across billions of cells.

#### Engineering zero-tolerance HARL compliance

Longevity gene editing demands velocity-engineered delivery: (1) Tissue-restricted LNPs with hepatocyte, cardiomyocyte, or neuronal tropism to limit the spatial forcing footprint. (2) Inducible editing systems (e.g. anti-CRISPR proteins, small-molecule-gated Cas variants) that allow fractionated editing: 5–10% of target cells per session over multiple treatments, keeping instantaneous editing velocity below the repair-capacity boundary. (3) Base editors and prime editors that avoid DSBs entirely, eliminating the most dangerous velocity-dependent toxicity (chromosomal rearrangements from simultaneous breaks). (4) Synthetic circuit controllers with negative-feedback: if edited-gene product exceeds physiological range, the circuit suppresses further expression—engineering σ_max into the construct itself. The principle is clear: longevity editing must be slow, fractionated, and reversibly gated—the opposite of oncology’s maximum-intensity paradigm.

### Panel 3 (Web Appendix): Real-time HARL surveillance—sensors, biomarkers, and the Z-score alarm architecture

The clinical translation of HARL requires continuous or high-frequency monitoring of the stress gap σ(t) and its first derivative dσ/dt. We propose a Z-score alarm architecture: each biomarker velocity v_X(t) is normalised against the patient-specific or population-derived HARL boundary, yielding a dimensionless alarm score Z_X(t) = v_X(t) / v_max,X. Values exceeding Z = 1.0 indicate HARL boundary approach; Z > 2.0 signals imminent breach requiring intervention. Multi-channel Z-scores are combined into a composite HARL Alarm Index (HAI) weighted by cascade position.

#### Tier 1: Continuous wearable sensors (seconds–minutes resolution)

- Heart rate variability (HRV): Autonomic adaptation velocity. Decline in SDNN or RMSSD >30%/hour signals baroreflex decompensation (τ_baro ≈ minutes). Direct readout of cardiovascular σ(t) for nifedipine-class perturbations. Devices: Apple Watch, Garmin, WHOOP, Oura Ring.
- Continuous core body temperature: Fever velocity (dT/dt in °C/h) is the earliest CRS signal. A rise >0.5°C/h predicts grade ≥2 CRS with ∼80% sensitivity. Ingestible thermometry (e.tempus, CorTemp) provides minute-level resolution; skin-surface patches (TempTraq, Fever Scout) offer non-invasive continuous monitoring.
- Continuous blood pressure: Beat-to-beat BP via cuffless photoplethysmography (Aktiia, Samsung Galaxy Watch BP) enables real-time dBP/dt computation. Velocity of BP drop during vasodilator therapy or BP rise during ESA correction can be mapped directly to the HARL corridor.
- Electrodermal activity (EDA) and skin conductance: Sympathetic activation velocity as a surrogate for autonomic stress gap. Rapid EDA increases (>2 μS/min) correlate with histamine release dynamics relevant to infusion reactions. Devices: Empatica E4/EmbracePlus.
- Respiratory rate and SpO2: Continuous pulse oximetry (Masimo, Nonin) captures the velocity of oxygen desaturation—a downstream indicator of pulmonary capillary leak in CRS. dSpO2/dt < −1%/h triggers the pulmonary HARL alarm.

#### Tier 2: Point-of-care biomarkers (minutes–hours resolution)

- Rapid ferritin assays: Immunoturbidimetric POC ferritin (Samsung LABGEO IB10, Siemens Atellica VTLi) returns results in 10–15 minutes, enabling 4–6-hourly velocity computation. Ferritin velocity >232 ng/mL per day as the primary macrophage HARL alarm.
- D-dimer POC: Fluorescence immunoassay (Radiometer AQT90 FLEX, Roche cobas h 232) with 12-min turnaround. D-dimer velocity >1.21 mg/L/day as the endothelial HARL alarm. Combined ferritin + D-dimer Z-scores form the two-stage cascade predictor validated in this manuscript.
- Point-of-care IL-6: Lateral flow and electrochemical IL-6 assays (Meso Scale Discovery, Ella microfluidic ELISA) achieving pg/mL sensitivity in <30 min. IL-6 velocity is the most direct cytokine cascade readout but lags macrophage activation by 12–24 h.
- Lactate: Capillary or interstitial lactate (Nova StatStrip, Lactate Scout) as a perfusion HARL alarm. Rising lactate velocity (>0.5 mmol/L/h) signals tissue hypoperfusion—the downstream consequence of sustained σ breach.
- Procalcitonin (PCT): Rapid PCT (bioMérieux VIDAS, Roche Elecsys) differentiates CRS from infection. PCT velocity aids differential diagnosis: infectious forcing follows different kinetics than immune-mediated CRS.

#### Tier 3: Continuous implantable and interstitial sensors (emerging)

- Continuous glucose monitors (CGM) repurposed: Interstitial glucose velocity during steroid-treated CRS; dextrose consumption rate as metabolic stress surrogate. Existing platforms (Dexcom G7, Abbott Libre 3) with 1-minute resolution.
- Implantable cytokine sensors: Electrochemical aptamer-based sensors for continuous IL-6, TNF-α, IFN-γ monitoring (research stage; Arroyo BioSciences, Nutromics patch technology). Would enable true real-time d[cytokine]/dt computation.
- Microdialysis catheters: Continuous interstitial sampling of metabolites and proteins. Research platforms (CMA Microdialysis) already used in neurocritical care; adaptation for subcutaneous cytokine monitoring in CRS is technically feasible.
- Smart IV line sensors: Inline spectrophotometric monitoring of infusion-related biomarkers (turbidity, haemolysis index) during vancomycin or bispecific antibody administration, enabling real-time infusion velocity adjustment.

#### The HARL Alarm Index (HAI): computational integration

The composite alarm integrates multi-channel Z-scores: HAI(t) = Σ w_i. Z_i(t), where weights w_i reflect each biomarker’s position in the pathological cascade (upstream signals weighted more heavily for early warning). Traffic-light output: Green (HAI < 1.0, all channels within HARL corridor), Amber (1.0 ≤ HAI < 2.0, one or more channels approaching boundary—increase monitoring frequency, alert clinical team), Red (HAI ≥ 2.0, HARL boundary breach imminent or active—initiate braking intervention). Braking interventions scaled to HAI magnitude: tocilizumab (IL-6 axis brake), corticosteroids (broad immune deceleration), infusion rate reduction (direct velocity control), vasopressors (haemodynamic support during adaptation lag), fractionated dosing hold (pause and reassess before next dose).

The engineering goal is a closed-loop HARL controller: continuous sensing → real-time Z-score computation → automated alarm escalation → protocolised braking response. This architecture transforms HARL from a theoretical framework into an implementable clinical decision support system, applicable across all velocity-governed therapeutic domains identified in this manuscript.

The foundational nifedipine study involved eight volunteers, though the prediction was validated in ∼8350 patients. The CAR-T analysis is retrospective; prospective interventional trials testing velocity-triggered intervention are needed. Current HARL uses population-average τ and σ^max^; patient-specific adaptive capacity biomarkers remain to be developed. Multi-system interactions and reversibility thresholds require further investigation. The D-dimer velocity data were derived from the longitudinal dataset with variable measurement intervals; standardised sampling protocols would improve precision.

The body tolerates destinations when it can accommodate the journey. Precision medicine and precision dosing have transformed therapeutic targeting, yet PK/PD theory remains fundamentally two-dimensional: how much and how long. The third dimension—the rate at which biological systems can adapt to pharmacologically driven state changes—has received insufficient formal attention, resulting in avoidable toxicity across therapeutic domains. HARL formalises this missing constraint. A single equation—|dS/dt| ^max^ = σ^max^/τ—unifies rate-dependent toxicity demonstrated prospectively four decades ago and independently confirmed across five therapeutic domains spanning minutes to weeks. Incorporating velocity governance into drug development offers three immediate opportunities: first, optimising the therapeutic index of toxic targeted therapies in oncology and haematology, where rate-aware dosing can widen the corridor between efficacy and tolerability; second, enabling earlier, rate-aware tolerability characterisation for innovative biological therapeutics—CAR-T constructs, bispecific engagers, mRNA platforms, and in-vivo gene editing—by characterising Maximum Tolerated Dynamics alongside Maximum Tolerated Dose from Phase I onward; and third, improving outcomes with existing approved therapies through velocity-guided re-optimisation of titration protocols. The transition from two-dimensional to three-dimensional PK/PD will require new clinical trial designs that incorporate velocity-escalation arms, new regulatory endpoints that specify allowable rates of change, and new monitoring architectures that track trajectories rather than snapshots. The goal is rate-aware medicine: therapies engineered to respect the third dimension from inception.

## Data Availability

Data sharing: Patient-level velocity data, longitudinal biomarkers, and Mathematica QSP source code are in the web
appendix. The Wei et al cohort is publicly available (ref 37)

## Acknowledgments

We thank the late John Urquhart for foundational discussions on rate-governed pharmacodynamics during the nifedipine programme (1984–1987).

## Methods

### QSP model and biomarker rationale

An 8-ODE model captured CAR-T expansion (dormant C^D^, engaged C^T^, memory C^M^), tumour dynamics (T, T^n^), and macrophage-cytokine signalling (M^a^, M^i^, IL-6). Three macrophage activation pathways— bystander, autocrine, kill-dependent—were parameterised by Monte Carlo optimisation against longitudinal IL-6 data from 15 individually-modelled patients across four cohorts (web appendix pp 3–8).^36^ The model identified ferritin velocity (macrophage readout) and D-dimer velocity (endothelial readout) as the biomarkers most directly coupled to the cascade (figure 5).

### Velocity calculation

For each patient and biomarker X, discrete slopes were computed over consecutive measurement pairs: v^X,i^ = (X^i+1^ − X^i^)/(t^i+1^ − t^i^). Early velocity = maximum positive slope. For ferritin, the early window (days 0–5) was used; for D-dimer, the same days 0–5 window captured the onset of endothelial activation. Two-stage thresholds by Youden-optimal grid search: ferritin >232 ng/mL per day (macrophage activation); D-dimer >1.21 mg/L per day (endothelial activation). A combined log-score (log^10^[ferritin velocity] + log^10^[D-dimer velocity]) was used for ROC analysis.

### Web Appendix

**Supplementary Table 1:** The velocity ledger — HARL parameters across therapeutic domains, with measured velocities and the dimensionless mismatch M.

| Domain | Adaptation $\tau$ | Reserve $\sigma$ | Boundary velocity $\sigma/\tau$ | Measured velocity $v$ |
| --- | --- | --- | --- | --- |
| Cardiovascular (nifedipine) | minutes | autonomic reserve | $\sim 0.2\text{--}0.3$ ng/mL/min | $\sim 2\text{--}3$ ng/mL/min |
| Haematology (ESA / CHOIR) | days-weeks | vascular compliance | $\leq 0.5$ g/dL per week | aggressive ($>$ boundary) |
| Radiation (DDREF) | 0.5–4 h | DNA-repair capacity | low dose-rate ($<0.1$ Gy/h) | $>1$ Gy/min |
| Infusion (vancomycin) | 2–10 min | mast-cell buffering | $\leq 10$ mg/min | rapid bolus ($>$ boundary) |
| Immunotherapy (CAR-T, N=202) | hours-days | immune regulation | 232 ng/mL/day (ferritin) | severe-CRS ($>$ boundary) |

Boundary velocity $\sigma/\tau$ from stated guideline limits and study rates; measured velocity from the same sources. No outcome or cost is shown — those belong to the fourth-dimension companion paper.

## Supplementary Material

## Appendix A: Mathematical derivation

From dA/dt = (S−A)/τ under constant forcing v = dS/dt, the steady-state gap is σ^ss^ = v.τ. Requiring σ^ss^ ≤ σ^max^ gives v ≤ σ^max^/τ. For arbitrary forcing, the convolution integral:

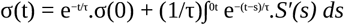

The supremum is bounded by τ.sup|S′(t)|, confirming the velocity constraint for arbitrary input waveforms. For periodic forcing with frequency ω, the amplitude transfer function |H(jω)| = 1/√(1+ω^2^τ^2^) shows that high-frequency components are attenuated by 1/(ωτ), providing the frequency-domain equivalent of the time-domain HARL bound.

## Appendix B: QSP model specification

The 8-ODE system:

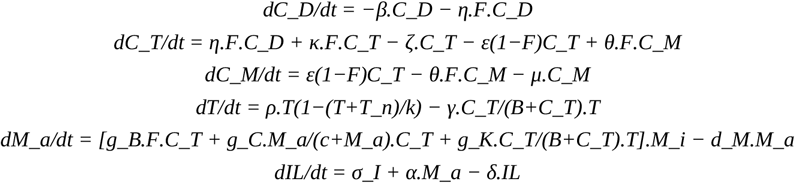

Parameters estimated by Monte Carlo (10 000 iterations/patient) with weighted residuals. Six macrophage model variants (K, B, KB, KC, BC, KBC) compared by AIC. Complete source: web appendix data files (master.m).

## Appendix C: Sensitivity analyses

The Velocity thresholds: ferritin >232 ng/mL per day, D-dimer >1.21 mg/L per day. Two operating modes: (1) safety rule-out—sensitivity 100%, specificity ∼78%; (2) Youden-optimised—sensitivity 91.1% (95% CI 79.4–97.2), specificity 93.6% (95% CI 88.9–96.9), AUC 0.95 (95% CI 0.91–0.98). Leave-one-out cross-validation confirmed robustness. Bootstrap stability (1000 resamples): thresholds stable within ±15%. Subgroup analyses: ALL and NHL subgroups yielded comparable discrimination. Subgroup analyses: ALL patients AUC 0.89; NHL 0.86; high tumour burden 0.90. The ferritin threshold showed sensitivity to measurement interval: 12-h sampling would shift the optimal threshold. Standardised protocols with fixed sampling times would reduce this source of variability.

## Appendix D: Clinical implementation protocol

1. Baseline (Day 0): Ferritin, D-dimer, IL-6, CRP, CBC, fibrinogen
2. Daily monitoring (Days 1–7): Repeat panel; standardise timing (±2 h)
3. Velocity calculation: Max positive slope over consecutive pairs in days 0–5
4. Risk stratification: Ferritin >232 ng/mL per day AND D-dimer >1.21 mg/L per day (dual-velocity gate)
5. Intervention: High-risk → early tocilizumab ± corticosteroids; ICU standby
6. Reassessment: Continue until velocity below threshold ≥48 h
7. Documentation: Velocity values, risk class, interventions in EMR

**Data files**: wei_cohort_processed.xlsx (patient-level data); master.m (Mathematica QSP code).

